# Autonomic Dysfunction in Gastroduodenal Disorders Evaluated through Multimodal Non-Invasive Physiological Testing

**DOI:** 10.1101/2025.06.29.25330513

**Authors:** Chris Varghese, Wendy Zhou, Armen Gharibans, Gabriel Schamberg, Sibylle Van Hove, Greg O’Grady, Leila Neshatian, Linda Nguyen

**Author notes:** **Corresponding Author** Prof Linda Nguyen, Stanford University, 430 Broadway Street, 3rd Floor, 94063 Redwood City, CA.

## Abstract

**Background:** Chronic gastroduodenal disorders are highly prevalent and encompass heterogeneous pathophysiology. Autonomic dysfunction may contribute, but its relative contribution to disease pathophysiology and symptoms is unclear. This study aimed to clarify the contributions of autonomic dysfunction in gastroduodenal disorders and symptom associations using multimodal testing, hypothesizing that distinct disorder subgroups would be revealed.

**Methods:** This prospective observational cohort study included 80 patients with chronic gastroduodenal symptoms. Participants underwent expert clinician-led autonomic evaluation, body surface gastric mapping (BSGM; Gastric Alimetry®, New Zealand), and gastric emptying scintigraphy testing. BSGM phenotyping, gastric emptying cut-offs, and autonomic criteria followed standard criteria.

**Results:** Gastroparesis was diagnosed in 27 (33.8%) patients. Autonomic dysfunction was present in 21 (26.3%) patients, and abnormal BSGM in 20 (25%). Autonomic dysfunction and abnormal BSGM were both more prevalent in gastroparesis (51.9% vs. 13.2%, p=0.001; and 44.4% vs. 15.1%, p=0.005, respectively). However, these diagnoses tended not to overlap, with patients having autonomic dysfunction generally exhibiting normal BSGM tests. Autonomic dysfunction independently predicted delayed gastric emptying (OR 13.8, p<0.001) and was associated with earlier meal responses (higher ‘meal response ratio’, p=0.04) and worse postprandial distress symptoms (p<0.05).

**Conclusions:** Autonomic dysfunction independently predicted delayed gastric emptying and revealed a gastroparesis mechanism distinct from gastric myoelectrical impairment. Autonomic dysfunction altered the postprandial gastric motor response and was associated with a greater symptom burden. Multimodal physiological testing including autonomic profiling reveals distinct mechanistic subgroups in patients with chronic gastroduodenal symptoms.

## Introduction

Chronic gastroduodenal disorders, encompassing gastroparesis, chronic nausea and vomiting syndromes (CNVS), and functional dyspepsia (FD), afflict >10% of the global population with significant impacts to patients and healthcare systems.^1–3^ The heterogenous pathophysiology underlying symptoms in these disorders makes targeted treatments challenging. The autonomic nervous system (ANS) plays a key role in integrating and regulating gastrointestinal (GI) tract functions. Autonomic dysfunction is therefore an important contributor to gastroduodenal pathophysiology,^4^ yet appears relatively under-investigated.

Dysfunctions in both vagal and sympathetic systems have important implications for GI sensory and motility functions, including modulating gastric emptying.^4,5^ For example, sympathetic dysfunction is described in up to 90% of patients with cyclic vomiting syndrome,^6^ and vagal dysfunction has been associated with FD pathogenesis.^7^ Further, the ANS has multiple important roles in co-regulating gastroduodenal function including for accommodation,^8^ gastric myoelectrical activity and motility,^9^ afferent satiety and immune signalling,^10^ antropyloric coordination,^11^ and pyloric sphincter tone.^12^ As such, autonomic impairments, have substantial potential to disrupt gastroduodenal physiology and generate chronic symptoms.

To date, gastric emptying testing has been the most widely used test of gastroduodenal function. However, its utility is limited by significant temporal variability, inconsistent symptom correlations, and a lack of sensitivity and specificity for both autonomic abnormalities and gastric myoelectrical dysfunction.^13,14^ We hypothesised that distinct pathophysiological subgroups underlie these heterogenous disorders that could be separated by multimodal physiological profiling. Besides autonomic testing, which is well established in clinical practice, body surface gastric mapping (BSGM) is a new test of gastric function that integrates time-of-test symptom profiling and non-invasive biomarkers of gastric myoelectrical function.^15,16^ This study therefore applied a multimodal physiological testing approach using gastric emptying testing, expert clinician-guided autonomic evaluation, and BSGM to define the relative contributions of autonomic dysfunction and gastric myoelectrical abnormalities to gastric physiological abnormalities and their symptom associations.

## Methods

This was a prospective observational cohort study conducted at Stanford Health, United States with institutional review and approval. The study is reported in accordance with the STROBE statement.^17^

### Eligibility criteria

Consecutive patients aged ≥18 years with chronic gastroduodenal symptoms and negative upper gastrointestinal endoscopy referred for specialist gastroenterology work-up at our tertiary center were eligible for inclusion. Exclusion criteria included those with known structural gastrointestinal diseases and previous abdominal surgery. Patients with cyclic vomiting syndrome or cannabinoid hyperemesis were also excluded. Specific exclusion criteria for Gastric Alimetry including BMI of >35, active abdominal wounds or abrasions, fragile skin, and allergies to adhesives were also applied.

### Autonomic testing

All patients were evaluated by an experienced autonomic neurologist. Those patients with symptoms and physical examination findings indicating autonomic dysfunction were referred for formal testing.^18^ All anticholinergics (including antidepressants, antihistamines, cough medications), sympathomimetics, parasympathomimetics, alpha and beta agonists, marijuana, and analgesics were withheld for 3 days prior to testing. Twelve hours prior to testing, patients were asked to avoid caffeine, alcohol, and nicotine.^19^ Postural orthostatic tachycardia syndrome was diagnosed if there was a sustained heart rate increase of 30 beats per minute or more within 10 minutes of head-up tilt without orthostatic hypotension. Quantitative sudomotor axonal reflex testing as described elsewhere.^19^ Cardiovagal testing included heart rate variability with deep breathing, and Valsalva ratio. Adrenergic testing included waveform analysis during the Valsalva. Autonomic specialists thereafter synthesised the following diagnostic categories, autonomic dysfunction, parasympathetic dysfunction, sympathetic dysfunction, and small fiber neuropathy.^19^

### Gastric Emptying Testing

All patients also underwent gastric emptying scintigraphy testing using standardized ∼255 kCal low-fat egg meal radiolabeled with ^99m^Tc. Delayed gastric emptying was present if gastric retention was >10% at 4 h.^20^ Gastroparesis diagnosis was confirmed based on cardinal symptomatology and delayed gastric emptying criteria.

### BSGM Testing

All patients also underwent BSGM using the Gastric Alimetry system (Alimetry, New Zealand), using a standardized protocol.^16^ Testing was performed after an overnight fast, and medications affecting GI motility were withheld for 48 h prior to testing. Patients were asked to avoid caffeine, nicotine, opiates, and cannabis on the day of testing. Glucose levels were controlled in diabetic subjects.

Briefly, the Gastric Alimetry system includes a high-resolution stretchable electrode array (8×8 electrodes; 20 mm inter-electrode spacing; 196 cm^2^), a wearable Reader, an iPadOS App for concurrent validated symptom logging during the test.^21–23^ Array placement was preceded by shaving if necessary, and skin preparation (NuPrep; Weaver & Co, CO, USA). Recordings encompassing 30 min fasting baseline, 10 min meal, and 4 h postprandial recording. Patients sat in a reclined relaxed position with limited movement, then transferred to the nuclear medicine table for imaging, with motion artifacts automatically corrected or rejected using validated algorithms.^24^ Symptoms including early satiation, nausea, bloating, upper gut pain, heartburn, stomach burn, and excessive fullness were measured continuously during testing at 15-minute intervals using 0-10 visual analog scales (0 indicating no symptoms; 10 indicating the worst imaginable extent of symptoms) and combined to form a ‘Total Symptom Burden Score’.^23^

BSGM phenotypes were determined on the basis of normative reference intervals for the following BSGM spectral metrics:^15^

- Gastric Alimetry Rhythm Index (GA-RI; reference interval: ≥0.25): a measure of rhythm stability calculated as the percentage of power contained in a narrow frequency band in the gastric range in an averaged spectrum, with adjustment for BMI, and reported as a scaled metric between 0 and 1.
- Principal Gastric Frequency (PGF; reference interval: 2.65-3.35 cycles per minute): slow wave frequency calculated as the frequency associated with the most stable oscillations at the body surface, determined by the GA-RI.
- BMI-adjusted amplitude (reference interval: 22-70 µV):^15^ measured amplitude at the body surface adjusted for individual’s BMI.

The ‘Meal Response Ratio’ (MRR) was also used to characterise meal response timing, which is calculated as the ratio of the average amplitude in the first 2 hours postprandially to that of the last 2 hours.^25^ MRR was not calculated if postprandial recording duration was <4 h. A normal MRR was empirically defined as >1 based on previous studies,^13,15,26^ meaning that the dominant gastric motor response should typically occur within the first two hours after a meal.

### Data analysis

All analyses were performed in R v.4.4.2 (R Foundation for Statistical Computing, Vienna, Austria). Continuous data were summarized as median (interquartile range) and compared by Mann-Whitney U test and Kruskal-Wallis tests as appropriate. Categorical data were cross-tabulated, presented as n (%), and differences tested using χ^2^ tests. Logistic regressions were performed to assess associations between physiological biomarkers and risk of delayed emptying, with results presented as odds ratios (OR) and 95% confidence intervals (CI).

## Results

Of 80 patients (median age 40 years, IQR 28-52; 79% female; median BMI 24.6, IQR 24.6-29.6) that were recruited, 27 (33.8%) had gastroparesis. Cohort demographics and diagnoses are detailed in **Table 1**. As per the methods, all patients were formally evaluated for signs and symptoms of autonomic impairments by a clinical specialist; of these 37 (46.3%) continued to formal quantitative autonomic testing, of whom 21/37 (56.8% total) demonstrated confirmed evidence of autonomic dysfunction. All patients also underwent Gastric Alimetry BSGM with 25% demonstrating spectral abnormalities, comprising low rhythm stability (n=10; 12.5%), low BMI-adjusted amplitude (n=2; 2.5%); and high frequencies (n=8; 10.0%).

**Table 1:** Demographics and results of Gastric Alimetry and autonomic testing stratified by gastric emptying status.

|  |  | **Normal GE** | **Gastroparesis** | **Total** | **p** |
| --- | --- | --- | --- | --- | --- |
| Age | Median (IQR) | 41.0 (28.0 to 50.0) | 32.0 (25.5 to 64.5) | 39.5 (27.5 to 52.2) | 0.819 |
| Sex | Female | 37 (69.8) | 26 (96.3) | 63 (78.8) | 0.014 |
|  | Male | 16 (30.2) | 1 (3.7) | 17 (21.2) |  |
| BMI | Median (IQR) | 25.2 (21.0 to 30.1) | 24.3 (19.9 to 29.0) | 24.6 (20.6 to 29.6) | 0.531 |
| GA-RI | Median (IQR) | 0.5 (0.3 to 0.7) | 0.5 (0.3 to 0.7) | 0.5 (0.3 to 0.7) | 0.76 |
| BMI-Adjusted Amplitude | Median (IQR) | 38.3 (30.6 to 52.5) | 34.9 (27.7 to 51.6) | 36.4 (28.6 to 52.7) | 0.427 |
| PGF | Median (IQR) | 3.1 (2.9 to 3.2) | 3.1 (3.0 to 3.4) | 3.1 (2.9 to 3.3) | 0.089 |
| Gastric Alimetry Phenotype Group | Low rhythm stability | 6 (11.3) | 4 (14.8) | 10 (12.5) | 0.005 |
|  | High frequency | 1 (1.9) | 7 (25.9) | 8 (10.0) |  |
|  | Low amplitude | 1 (1.9) | 1 (3.7) | 2 (2.5) |  |
|  | Normal spectral metrics | 45 (84.9) | 15 (55.6) | 60 (75.0) |  |
| Autonomic Dysfunction | No | 46 (86.8) | 13 (48.1) | 59 (73.8) | 0.001 |
|  | Yes | 7 (13.2) | 14 (51.9) | 21 (26.2) |  |
| Parasympathetic Dysfunction | No | 5 (62.5) | 6 (42.9) | 11 (50.0) | 0.658 |
|  | Yes | 3 (37.5) | 8 (57.1) | 11 (50.0) |  |
| Sympathetic Dysfunction | No | 1 (12.5) | 7 (50.0) | 8 (36.4) | 0.194 |
|  | Yes | 7 (87.5) | 7 (50.0) | 14 (63.6) |  |
| Small Fiber Neuropathy | No | 51 (96.2) | 23 (85.2) | 74 (92.5) | 0.185 |
|  | Yes | 2 (3.8) | 4 (14.8) | 6 (7.5) |  |
| POTS | No | 47 (88.7) | 18 (66.7) | 65 (81.2) | 0.037 |
|  | Yes | 6 (11.3) | 9 (33.3) | 15 (18.8) |  |
| EDS | No | 44 (83.0) | 22 (81.5) | 66 (82.5) | 1.00 |
|  | Yes | 9 (17.0) | 5 (18.5) | 14 (17.5) |  |

BMI, body mass index; GA-RI, Gastric Alimetry Rhythm Index; PGF, Principal Gastric Frequency; POTS, postural orthostatic tachycardia syndrome; EDS, Ehlers-Danlos Syndrome; IQR, interquartile range; GE, gastric emptying.

### Gastroparesis phenotypes

Patients with gastroparesis, as confirmed on scintigraphy testing, were more likely to have impaired autonomic function (51.9% vs 13.2%; p=0.001) and were also more likely to have an abnormal Gastric Alimetry test (44.4% vs 15.1%; p=0.005; **Table 1** and **Figure 1**). However, distinct gastroparesis patient subgroups were apparent, such that autonomic dysfunction in gastroparesis predominantly occurred in patients with a normal Gastric Alimetry (normal in 80% vs abnormal in 16.7%; p=0.004; **Figure 2**).

**Figure 1:**
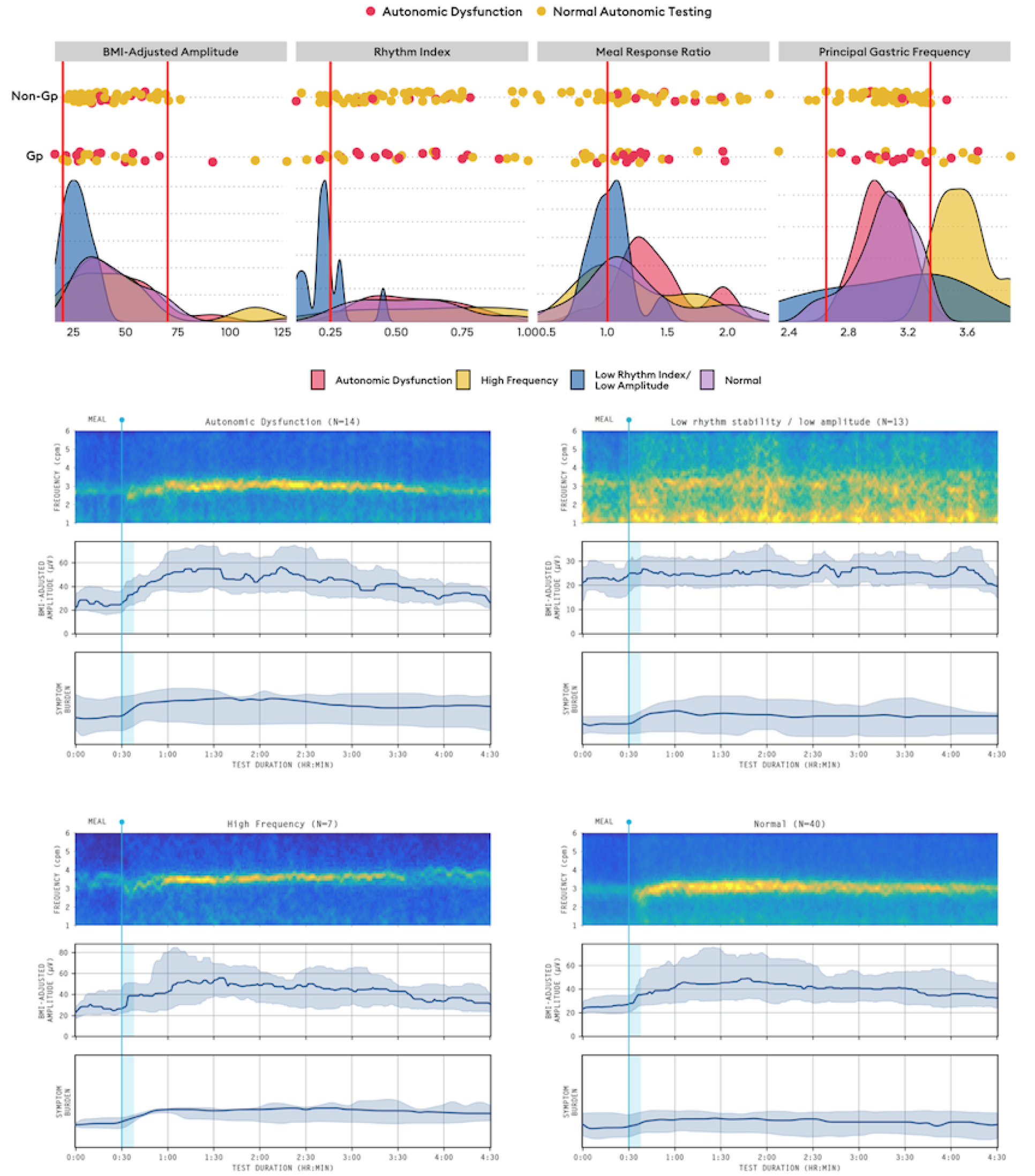
Scatter and density plot by Gastric Alimetry spectral metrics, stratified by emptying status and coloured by autonomic function status. Notable trends include autonomic dysfunction predominantly occurring within normal spectral metric reference intervals,^15^ and most individuals with autonomic dysfunction having an early meal response (based on a meal response ratio >1).^25^

**Figure 2:**
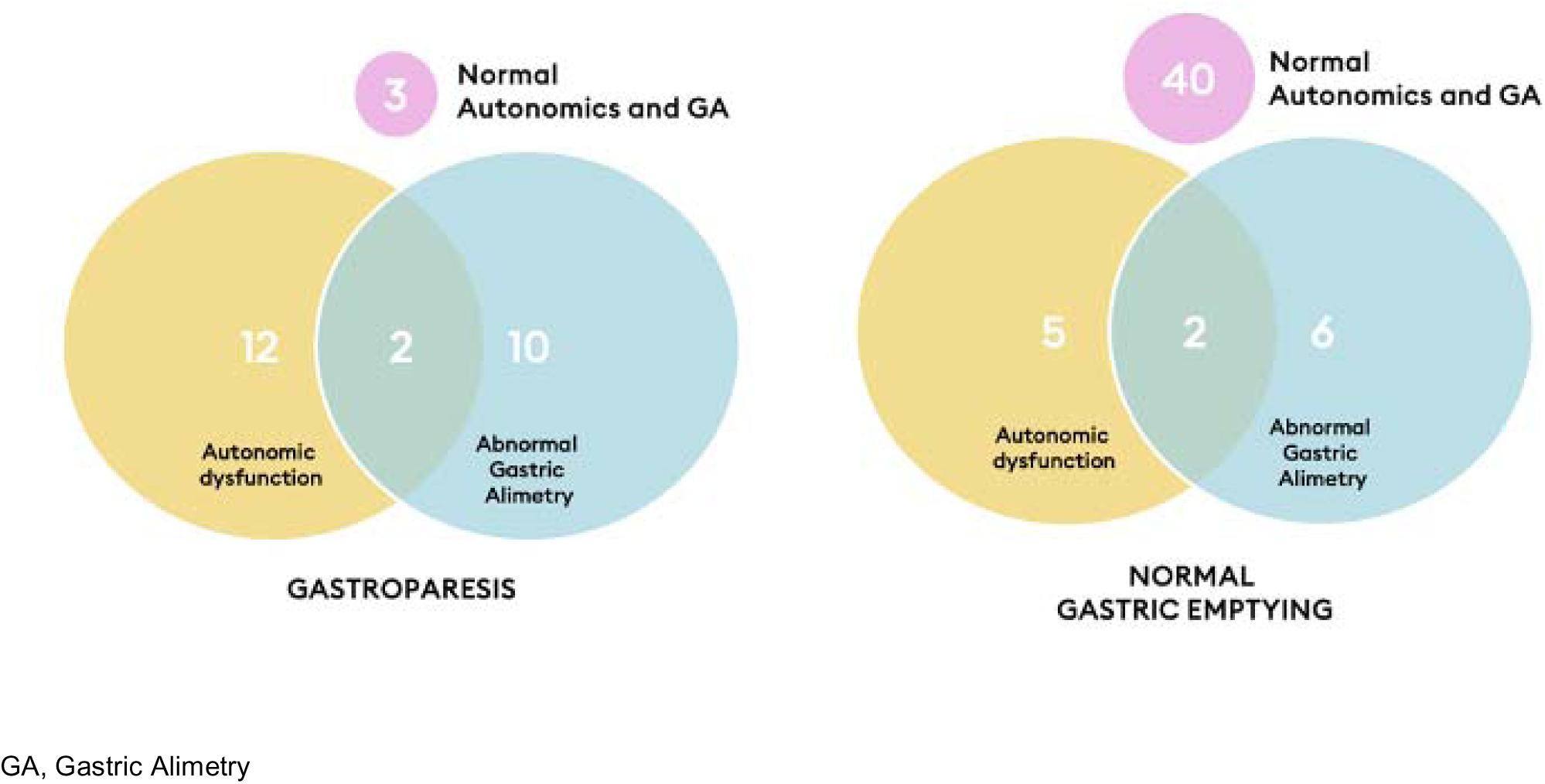
Distinct subgroups were apparent in gastroparesis; the majority of patients with gastroparesis and autonomic dysfunction had a normal Gastric Alimetry test (80% vs 16.7%; p=0.004). In patients without gastroparesis, the majority had both normal Gastric Alimetry and autonomic function.

Based on multimodal testing, the most common abnormalities in patients with gastroparesis were autonomic dysfunction (44.4%; n=12), high gastric frequencies (22.2%; n=6), and low rhythm stability and/or amplitudes (22.2%; n=6). Only 11.1% (n=3) of patients with gastroparesis had both normal BSGM and autonomic function, compared to 75.5% of patients with normal gastric emptying having both tests as normal. A physiological abnormality on multimodal testing was therefore highly predictive of delayed emptying (OR 24.6, 95% CI 7.2-116.2; p<0.001). Of all Gastric Alimetry spectral phenotypes, high frequency had the greatest association with gastroparesis (OR 28.13, 95% CI 3.78-606; p=0.005). Notably, autonomic dysfunction was also independently associated with delayed gastric emptying when accounting for gastric myoelectrical abnormalities detected on BSGM (OR 13.8, 95% CI 3.87-58.4; p<0.001; **Figure S1**).

### Effects of autonomic dysfunction on gastric motor function

The high majority of individuals with autonomic dysfunction had normal BSGM spectral metrics including rhythm stability, frequencies, and amplitudes (81%, n=17). However, autonomic dysfunction was associated with an altered meal response profile as evaluated by BSGM, showing a greater proportion of the gastric motor response occurring in the first 2 h postprandially (median MRR 1.3 [IQR 1.2-1.5] vs 1.1 [IQR 1.0-1.4], p=0.04; **Figure 3**). This difference was most pronounced in the subset of patients who also had small fiber neuropathy (median 1.7 [IQR 1.5-2.0] vs 1.2 [IQR 1.0-1.4], p=0.01; **Figure 3**). Gastric amplitude, frequency and rhythm were comparable between those with and without autonomic dysfunction, including individuals with POTS (p>0.1).

**Figure 3:**
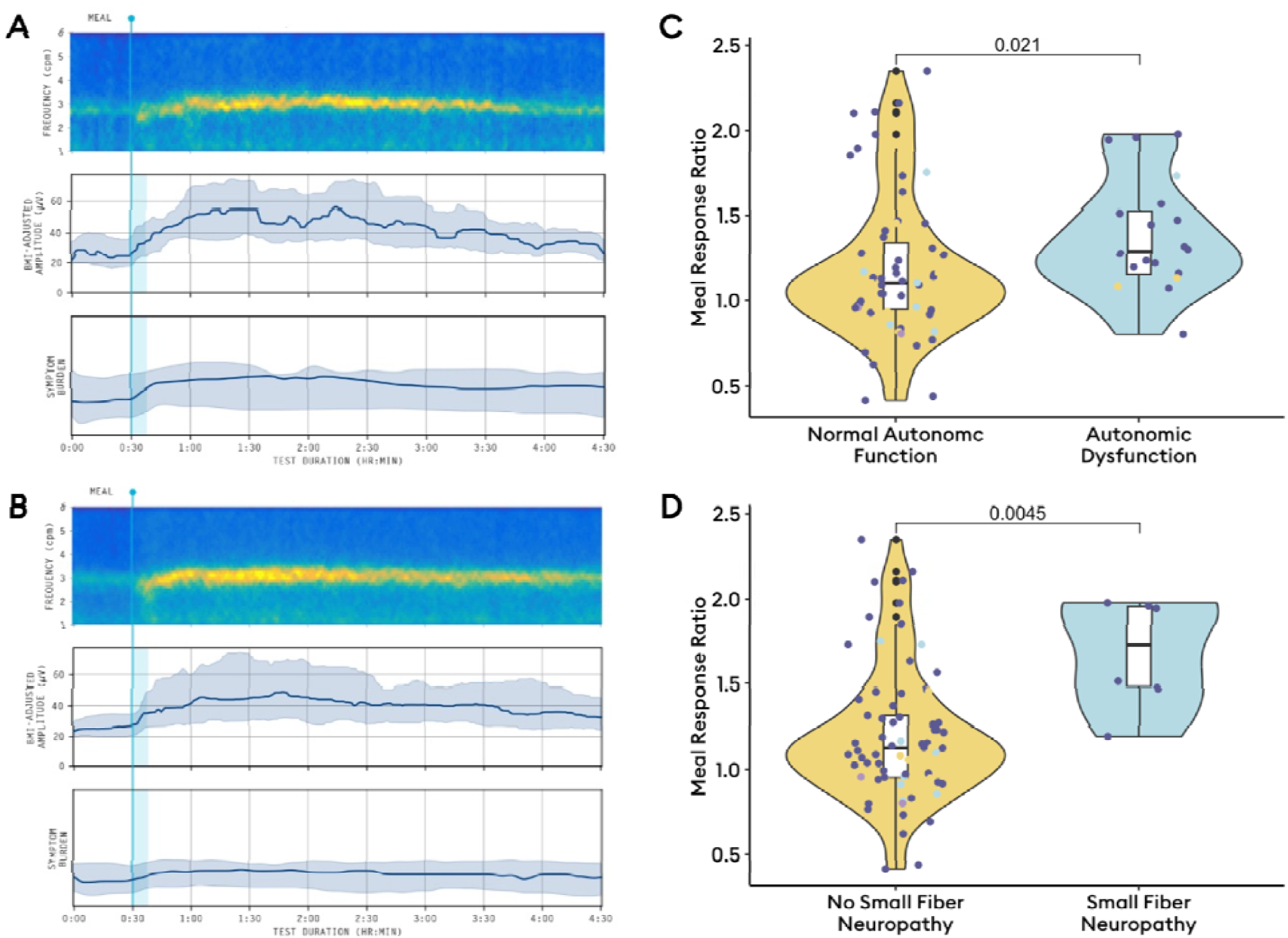
Average spectrograms, amplitude curves, and symptom profiles. A: normal autonomic function (median meal response ratio [MRR] 1.3); B: autonomic dysfunction (median MRR 1.1) evidenced by left-shifted meal responses and higher symptoms; C: box and violin plot comparing MRR by autonomic dysfunction; D: box and violin plot comparing MRR by presence of small fiber neuropathy.

Exploratory analysis of autonomic metrics from formal testing found a negative correlation between BMI-adjusted amplitude and the valsalva ratio (R = −0.36, p=0.04). There were also moderate-to-strong correlations between BMI-adjusted amplitude and baseline diastolic blood pressure and heart rate during tilt table testing (R = 0.37-0.71; p<0.05), and an inverse correlation with baseline systolic blood pressure (R = −0.37, p=0.04). However, these exploratory findings were not statistically significant after correction for multiple comparisons (**Figure S2**).

### Symptom associations with autonomic dysfunction

In the overall cohort, autonomic dysfunction was associated with higher heartburn scores (2.2 vs 1.0, p=0.04). When stratified by Gastric Alimetry results, the effect of autonomic dysfunction on symptoms differed. In those with abnormal Gastric Alimetry spectral analysis, there were no differences in symptoms by autonomic dysfunction. However, among patients with normal Gastric Alimetry, 28.3% (n=17) had autonomic dysfunction, and these patients experienced markedly worse postprandial distress (excessive fullness 4.5 vs 1.9, p=0.03; early satiation 5.0 vs 2.0, p=0.007), heartburn (1.3 vs 0, p=0.02), and total symptom burden (28.3 vs 13.8, p=0.06; **Figure 4**).

**Figure 4:**
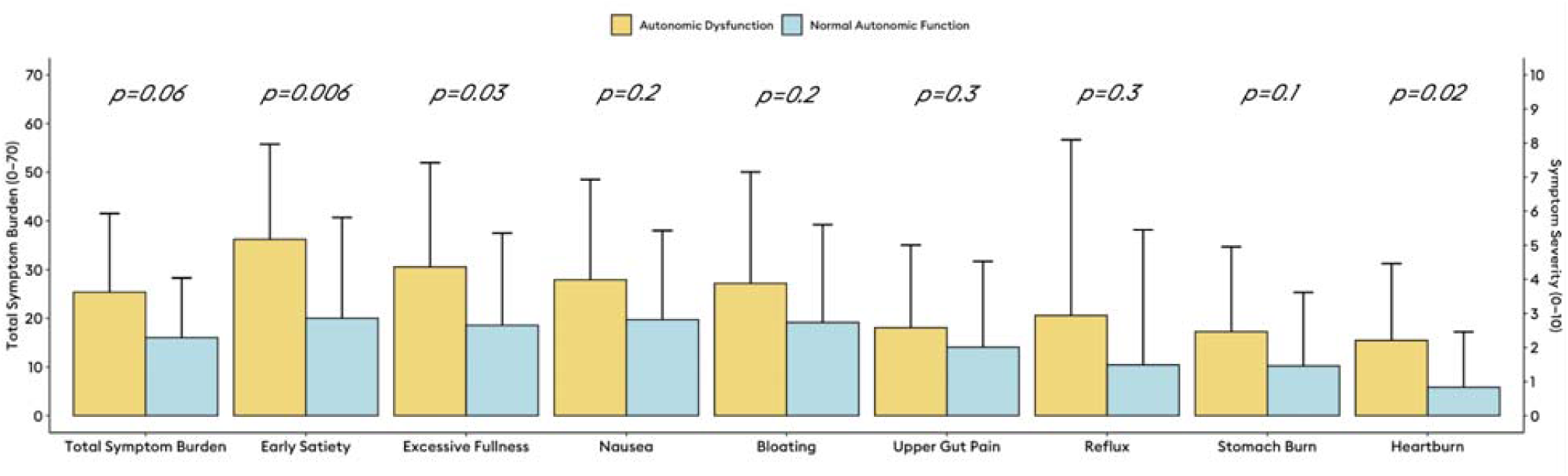
Symptom differences in patients with normal Gastric Alimetry stratified by presence of autonomic dysfunction. Data are presented as mean and standard deviation.

## Discussion

This study confirmed the hypothesis that multimodal physiological testing, including assessments of autonomic dysfunction, gastric transit, and gastric myoelectrical function, can reveal distinct pathophysiological subgroups among patients with chronic gastroduodenal symptoms. Notably, autonomic dysfunction contributed to delayed gastric emptying independent of gastric myoelectrical impairments, indicating that these tests capture distinct mechanisms underlying gastroparesis. Moreover, autonomic dysfunction was most commonly observed in patients with normal Gastric Alimetry spectral analysis where it was associated with altered gastric meal responses and worse postprandial distress symptoms.

Multimodal testing applied here enabled effective subtyping of a heterogenous cohort of patients. Our findings therefore reinforce and extend existing evidence emphasising the important role of autonomic dysfunction in gastroduodenal disorders,^4^ and suggest that it,alongside high-resolution myoelectrical recordings, should be more widely incorporated into the work-up of undifferentiated foregut symptoms. This mechanistic subgrouping is particularly helpful in delineating gastroparesis etiologies, as delayed transit can be caused by multiple physiological abnormalities including neuromuscular disorders, impaired fundic accommodation, and pyloric factors.^25,27^ Notably, vagal functions play a crucial role in regulating fundic accommodation, pyloric resistance,^28^ and the timing of the postprandial motor response, all of which impact gastric emptying.^25,29^

Autonomic dysfunctions vary, potentially leading to a range of gastrointestinal sensory and motor effects. This study showed that impaired autonomic regulation is linked to altered gastric meal responses and increased postprandial distress, while also presenting preliminary evidence that higher gastric amplitudes may reflect reduced autonomic flexibility. Together with prior findings that autonomic disturbances can result in both rapid and delayed gastric emptying,^5^ and that sympathetic hypofunction predominates in patients with nausea and vomiting symptoms irrespective of gastric emptying,^4^ these results underscore the complex interactions between autonomic regulation and gastroduodenal pathophysiology. These new insights should inform additional multimodal studies to confirm and extend these relationships in future.

In separate work, diabetic gastropathy has been linked to various myoelectrical abnormalities, including including elevated gastric frequencies in association with marked neuropathy,^30^ and loss of rhythm stability likely related to interstitial cells of Cajal depletion.^30–32^ High gastric frequencies have also been seen in the setting of surgical vagotomy after fundoplication, leading to suggestions this high frequency in general may be an emerging biomarker of gastric vagal injury.^33,34^ Previous studies have also described a progression of diabetic autonomic neuropathy that begins as parasympathetic hypofunction, followed by sympathetic excess and later sympathetic hypofunction with cardiovascular involvement.^35,36^ This trajectory may mirror the range of spectral phenotypes observed in these cohorts, from altered meal responses, to elevated gastric frequencies, and persistent dysrhythmia. Future work should further clarify the distinct roles of parasympathetic and sympathetic dysfunction in these gastric myoelectrical abnormalities to extend these observations.

The mechanistic role of autonomic dysfunction in these highly prevalent gastroduodenal disorders has important therapeutic implications. Of note, randomized studies have supported the efficacy of non-invasive vagal nerve stimulation in a range of gastrointestinal disorders,^37^ with recent evidence demonstrating normalisation of postprandial amplitudes, at sufficient stimulation frequencies in association with increased parasympathetic and reduced sympathetic tone.^38^ Similar effects have also been proposed using direct gastric neuromodulation.^39^ Effectively identifying patients with autonomic contributors to gastroduodenal symptoms, could therefore enable targeted selection for these novel treatments, whereas patients with gastric myoelectrical abnormalities may benefit more from direct therapies.^40^

One key limitation of this real world prospective series is that not all patients underwent formal autonomic testing. Nevertheless, the work-up was guided by expert clinicians in autonomic function assessment who based their decisions on standard clinical evaluations. Additionally, patients were referred for autonomic evaluation based on clinical suspicion for dysautonomia and did not include patients with isolated gastric symptoms. Our current cohort study was also not specifically powered to evaluate relationships between specific autonomic metrics, transit, and Gastric Alimetry test metrics. The findings of this study are nonetheless novel, and warrant larger studies with standardized autonomic testing to robustly define the autonomic contributors to gastroduodenal disorders, including evaluating the individual contributors of specific components of the ANS. When considering the generalisations of these findings, it is also worth noting that this study had a minority of patients with diabetes (n=6), such that alternative mechanisms for autonomic and gastric dysfunctions predominated in the study’s referral base.

In conclusion, autonomic dysfunction is an important disease mechanism in chronic gastroduodenal disorders and could be a driving mechanism in a large subset of gastroparesis patients, independent of gastric myoelectrical impairments. These findings underscore the need for comprehensive physiological evaluations that go beyond gastric emptying testing - including both autonomic function and gastric myoelectrical assessments - to more comprehensively subtype patients to inform personalized care.

## Data Availability

All data produced in the present study are available upon reasonable request to the authors.

## Acknowledgements

N/A

---

**Figure S1:**
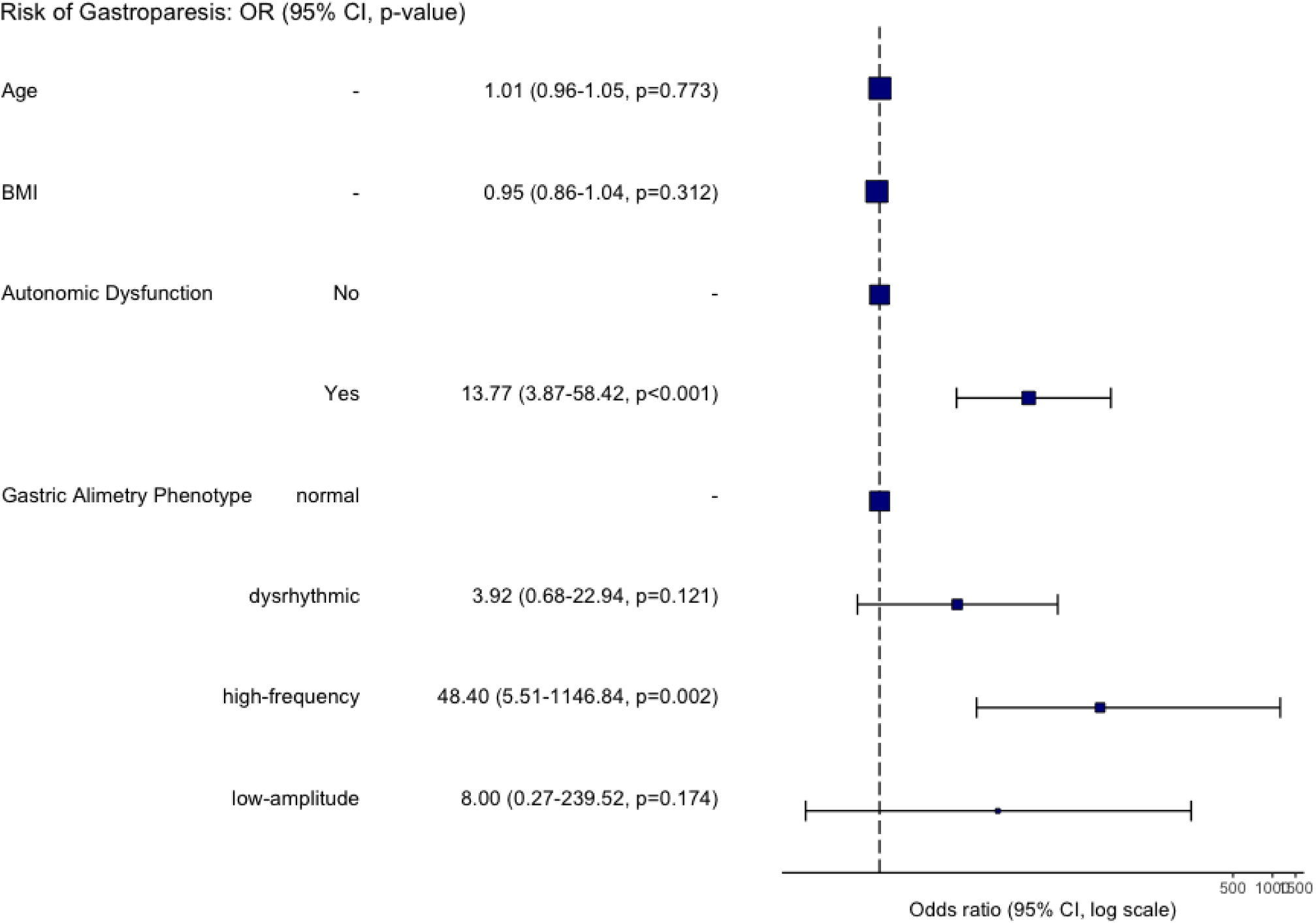
Logistic regression analysis of independent effects of autonomic dysfunction and Gastric Alimetry phenotypes on risk of delayed gastric emptying

**Figure S2:**
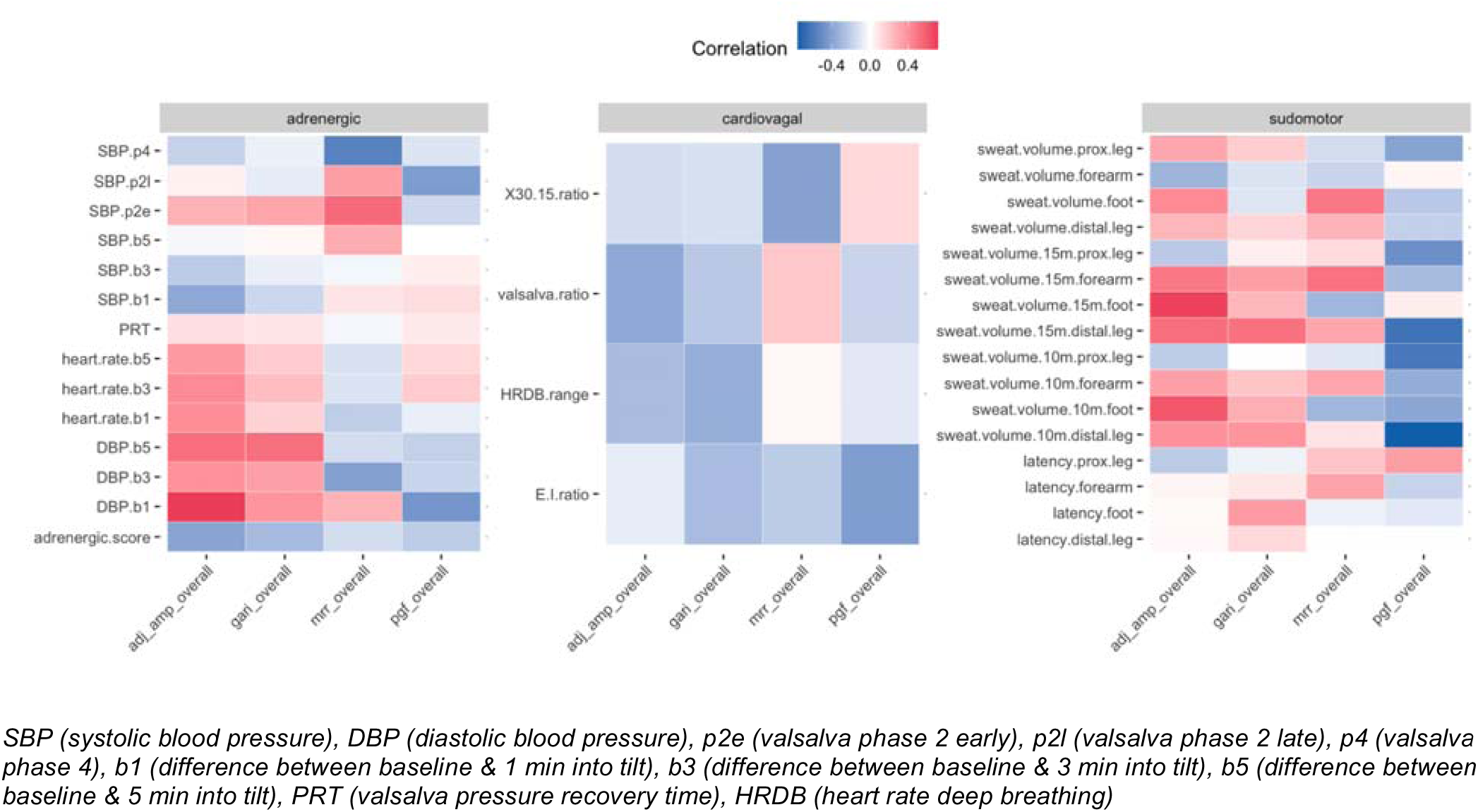
Exploratory analysis of correlations between autonomic testing metrics and Gastric Alimetry spectral metrics.

